# Instantaneous-Frequency EEG Microstate Dynamics Stratify Motor Subtypes in Parkinson’s Disease

**DOI:** 10.64898/2026.06.09.26355312

**Authors:** Sou Nobukawa, Hirohisa Watanabe

## Abstract

Parkinson’s disease (PD) is clinically heterogeneous, yet objective electrophysiological markers of its postural-instability/gait-difficulty (PIGD) and tremor-dominant (TD) motor subtypes are lacking. We tested whether the temporal dynamics of instantaneous-frequency (IF) microstates in resting-state electroencephalography (EEG) distinguish these subtypes from each other and from healthy controls (HC). In a publicly available cohort (OpenNeuro ds007526) comprising 28 HC and 97 PD patients classified as PIGD (n = 50) or TD (n = 47), the spatial distribution of the IF was reduced by principal component analysis and modeled with a Gaussian hidden Markov model, yielding three recurrent microstates. Per-participant mean dwell time, occupancy, and state-transition probabilities were compared across the three groups and, within PD, correlated with clinical scores. We found that the dynamics of one microstate varied systematically across groups: its dwell time, occupancy, and self-transition probability increased monotonically from HC through TD to PIGD, while outgoing transitions decreased, so that the state became an increasingly persistent attractor. For dwell time, all three pairwise contrasts survived correction (HC versus PIGD, Hedges’ g = 1.06; HC versus TD, g = 0.59; PIGD versus TD, g = 0.40). None of the dynamic indices was associated with clinical severity, disease duration, or medication dose within PD. IF-microstate dynamics thus stratify the PD motor subtypes along a graded continuum without tracking continuous disease severity. The approach offers a candidate objective EEG marker for motor-subtype stratification, complementing spectral characterizations of PD.

## I. Introduction

**P**arkinson’s disease (PD) is the second most prevalent age-related neurodegenerative disorder after Alzheimer’s disease, and its global burden has more than doubled over the past three decades, with continued rapid growth projected as populations age [1]. PD is clinically heterogeneous: patients are commonly stratified into the postural-instability and gait-difficulty (PIGD) and tremor-dominant (TD) motor subtypes [2], [3], which diverge markedly in long-term prognosis, with PIGD patients experiencing faster cognitive decline, greater non-motor burden, and a higher risk of dementia and falls than their TD counterparts [4]–[6]. Objective biomarkers capable of capturing this subtype divergence in clinically diagnosed patients—and, in the longer term, of detecting pre-motor neurodegeneration in at-risk and prodromal populations [7]— remain a central unmet need.

Electroencephalography (EEG) has long served as a non-invasive functional readout in the study of age-related neurodegenerative disorders, capturing electrocortical signatures of disease-related changes in brain network activity complementary to those obtained from structural imaging [8]–[10]. Its practical advantages—low cost, portability, and millisecond-scale temporal resolution—make it particularly well-suited to longitudinal monitoring and to integration with other physiological measurements in multimodal biomarker pipelines [11]. In PD specifically, the most extensively characterized EEG biomarkers are posterior alpha slowing and frontal theta enhancement [12], [13]. These are, however, time-averaged spectral measures that, by construction, cannot resolve the transient cortical dynamics through which subtype-dependent differences may be expressed.

Beyond signal amplitude, dynamic features derived from the phase component of cortical activity have increasingly been recognized as carrying complementary information about large-scale spatiotemporal organization, encompassing not only inter-regional synchrony but also the transient reconfiguration of spatial patterns and the dynamic structuring of brain networks [14]–[16]. Phase-based dynamic analyses have detected widespread alterations of functional network dynamics in healthy aging and in age-related neurodegenerative disorders such as Alzheimer’s disease [17], [18], and theta-band phase synchronization has been shown to underpin the hub-based network architecture that supports cognitive function in older adults [19]. Instantaneous frequency (IF), defined as the temporal derivative of the unwrapped phase, provides direct access to one facet of these phase-derived dynamics; the IF-microstate framework [20], [21] characterizes its multi-channel spatial distribution as a sequence of quasi-stable discrete states, thereby describing large-scale cortical activity through the lens of dynamic transitions among spatial patterns rather than through direct quantification of inter-regional synchrony or interaction. In parallel, the noradrenergic system is thought to modulate the balance between functional integration and segregation across cortical areas [22], with consequences that should manifest in the large-scale spatiotemporal organization of cortical activity. In PD, the noradrenergic system undergoes activity-level alterations—including compensatory hyperactivity of surviving LC neurons and downstream changes in cortical noradrenergic signalling [23], [24]—that are expected to perturb such large-scale cortical organization. IF-microstate analysis—by tracking the temporal organization of phase-derived activity across multiple channels—emerges as a candidate framework for characterizing subtype-related differences in large-scale cortical dynamics in PD; to our knowledge, however, its applicability to PD subtype differentiation has not been examined.

In this context, we hypothesized that applying IF-microstate analysis to resting-state EEG of PD patients would reveal a new aspect of alterations in large-scale cortical dynamics that complements the time-averaged spectral biomarkers established to date. To test this hypothesis, we analyzed the publicly available OpenNeuro ds007526 dataset [25], in which PD patients are stratified into PIGD and TD motor subtypes according to the Stebbins criteria computed from Movement Disorder Society Unified Parkinson’s Disease Rating Scale (MDS-UPDRS) scores [3]. Following standardized preprocessing of the resting-state recordings, IF-microstate sequences were extracted and decoded via a hidden Markov model, and dwell time, occupancy, and transition probabilities were compared across the HC, PIGD, and TD groups. The remainder of this paper is organized as follows: Section II reviews related work, Section III details the materials and methods, Section IV presents the results, and Section V discusses their implications.

## II. Related Works

### A. Motor Subtypes and Differential Neurodegeneration in Parkinson’s Disease

The clinical heterogeneity of PD has long motivated the use of motor sub-typing to capture biologically meaningful patient strata. Jankovic *et al*. [2] provided the foundational scheme by dividing the Deprenyl And Tocopherol Antioxidative Therapy Of Parkinsonism (DATATOP) cohort, a large early-PD trial cohort followed prospectively in North America, into PIGD and TD subtypes based on motor symptom profiles. Stebbins *et al*. [3] subsequently formalized this scheme for the MDS-UPDRS, introducing a tremor-dominance ratio computed from the motor sub-scores that operationally classifies patients into PIGD, TD, and indeterminate categories. The MDS-UPDRS-based scheme has since been widely adopted as a reproducible standard for subtype stratification in observational and biomarker studies, and underlies the subtype assignment used in the present work (Section III). Crucially, this classification has been shown to track not only the symptomatic profile but also distinct trajectories of underlying neurodegeneration [5], [6], the basis for which is summarized below.

At the pathological level, Braak *et al*. [26] described a stereotyped staging in which *α*-synuclein inclusions progress from the dorsal motor nucleus of the vagus through the LC before reaching the substantia nigra and cortex, indicating that early PD pathology is, in part, extra-nigral. The LC noradrenergic (LC-NA) system is one of the most consistently affected extra-nigral systems: post-mortem and neuromelanin-sensitive MRI studies document LC neuronal loss across the PD spectrum [27], [28], with comparable LC signal reduction already observable in prodromal REM sleep behavior disorder [29], and clinical correlations link LC integrity to both motor and non-motor symptom burden [30].

LC structural degeneration, however, does not translate one-to-one into cortical NA depletion. In animal models of PD, surviving LC neurons show compensatory increases in firing rate, alterations in NA release, and downstream sensitization of cortical adrenergic receptors [23], [24]; in vivo human PET/neuromelanin-MRI imaging finds LC neuromelanin signal and cortical *α*_2_-adrenergic receptor density only partially coupled [30]; and substantial tissue NA depletion is observed mainly at the PD-with-dementia stage [31]. Although noradrenergic deficiency has been linked to the PIGD phenotype [32], [33], PD-related noradrenergic alterations are not limited to NA depletion. Capturing this broader multi-level reorganization—spanning receptor sensitization, transmitter release, and large-scale cortical network dynamics— is therefore important for understanding the noradrenergic contribution to PD pathophysiology.

These pathological differences translate into substantial divergence in clinical trajectory. PIGD patients exhibit a faster rate of cognitive decline and a higher cumulative incidence of dementia than TD patients, as documented in long-term prospective follow-up of incident PD cohorts [4], [5] and further summarized in subsequent reviews [6]. Taken together, motor subtype constitutes a strong prognostic axis that meaningfully stratifies disease course, motivating the development of biomarkers that can resolve subtype-related differences in the underlying neural dynamics.

### B. Resting-State EEG Studies in Parkinson’s Disease

Among EEG biomarkers in PD, the most consistently reported features include global slowing of the spectrum, posterior alpha attenuation, and frontal theta/delta enhancement [13], [34]. This pattern was systematically described in an early quantitative EEG study by Soikkeli *et al*. [12] and has since been widely replicated across clinical centres and recording protocols, with greater slowing observed in patients exhibiting cognitive impairment [13], [35].

Spectral power summarizes frequency content averaged over the recording, whereas entropy- and complexity-based measures capture dynamic properties of the EEG signal that are not accessible from spectral analysis alone [36]. Sample entropy and the related approximate entropy [37], [38], multiscale entropy [39]–[41], and Lempel–Ziv complexity [42] have all been applied to characterize EEG in healthy aging and in age-related neurodegenerative disorders, revealing distinct patterns of entropy change that differentiate physiological aging, Alzheimer’s disease, and PD. In PD specifically, complexity measures have been related to cognitive status and have shown sensitivity to disease progression [36], providing a perspective on cortical activity that complements but does not overlap with spectral indices.

A complementary line of work has examined large-scale functional connectivity and network topology of resting-state EEG and magnetoencephalography (MEG) in PD [34], [43]. Phase-based connectivity measures such as the phase lag index (PLI) [44] and the weighted PLI [45], together with amplitude-based measures such as orthogonalized amplitude envelope correlation [46], have been used to characterize static inter-regional coordination. Graph-theoretic analyses of such connectivity matrices have reported network disruptions, particularly in patients with cognitive impairment [47]. A recent systematic review by Wang *et al*. [34] synthesizes the relationships between EEG-derived markers—spanning spectral, connectivity, and complexity indices—and both cognitive and motor symptoms in PD.

Conventional EEG microstate analysis, pioneered by Lehmann [48] and operationalized in its widely used four-state form by Koenig *et al*. [49], characterizes resting-state EEG as a sequence of quasi-stable topographies derived from the global field power and has been applied across a broad range of psychiatric and neurological conditions [50], [51]. While its application to PD remains comparatively limited, the framework has motivated growing interest in dynamic descriptions of cortical activity in this disorder. Two limitations are nevertheless shared by the body of work surveyed above. First, the majority of established dynamic indices—including conventional microstates—are derived from amplitude- or power-related properties of the EEG signal; while phase-based measures such as PLI and wPLI have been used to characterize inter-regional connectivity, the phase-based dynamic spatial reconfiguration of cortical activity—which IF-microstate analysis explicitly targets—remains largely uncharacterized in PD. Second, this methodological orientation has constrained subtype-related findings, a gap reflected in the limited subtype coverage of two recent systematic reviews [34], [43]. The IF-microstate framework, described next, opens this methodological space by providing a phase-based dynamic descriptor, and its application to PD subtype stratification in the present study contributes to closing the resulting empirical gap.

### C. Instantaneous-Frequency Microstate Analysis

The IF-microstate framework is built on the analytic-signal representation of multichannel EEG. After bandpass filtering to a frequency range of interest, the Hilbert transform yields, for each channel, an unwrapped phase whose temporal derivative defines IF. At every time point, the IF values across electrodes form a multidimensional vector capturing the spatial distribution of phase-derived dynamics, and clustering these vectors over time gives a sequence of quasi-stable IF microstates. The framework thus stands as a phase-based counterpart to the classical microstate framework, which segments EEG by the topography of global field power [48], [50].

This framework was first formally introduced by Nobukawa *et al*. [20], who applied *k*-means clustering to the spatial IF vectors and characterized the resulting state sequence by centroid topographies, state occupancies, and transition probabilities. In this *k*-means formulation, each time point is assigned to a state on the basis of spatial similarity to its centroid alone, and the dynamic indices are obtained post-hoc as descriptive statistics of the labeled sequence. Using this approach, atypical spatio-temporal patterns of neural dynamics were identified in Alzheimer’s disease, demonstrating that phase-derived spatial patterns carry information complementary to amplitude-based descriptors.

Nobukawa *et al*. [21] subsequently extended this framework by replacing the *k*-means stage with a hidden Markov model (HMM), in which the transition structure between states is incorporated as a constituent of the model rather than estimated post-hoc. With this extension, the state definitions themselves reflect both the spatial structure of IF topographies and the temporal continuity of state-to-state transitions. This extended framework revealed age-group differences in the spatiotemporal organization of resting-state EEG between younger and middle-aged adults. The present study extends this line of work by applying the IF-microstate–HMM framework to motor-subtype differentiation in PD.

## II. Materials and Methods

### A. Analysis Pipeline Overview

Figure 1 summarizes the overall analysis pipeline. Resting-state EEG recordings from the OpenNeuro ds007526 dataset are first stratified by Stebbins criteria into HC, PIGD, and TD groups (Section III-B). Each recording is then preprocessed using a standardized pipeline that includes line-noise removal, bandpass filtering, bad-channel handling, ICA-based artefact rejection, epoching, and automated epoch rejection (Section III-C). The cleaned epochs are passed to the IF-microstate–HMM analysis [Fig. 1(B)], in which a hidden Markov model is trained on the pooled data of all three groups to define a common state set, and per-subject state sequences are subsequently recovered by Viterbi decoding (Section III-D). From these sequences, dwell time, occupancy, and transition probabilities are computed at the subject level (Section III-E) and used for between-group comparison and within-PD clinical correlation analysis (Section III-F).

**Fig. 1.**
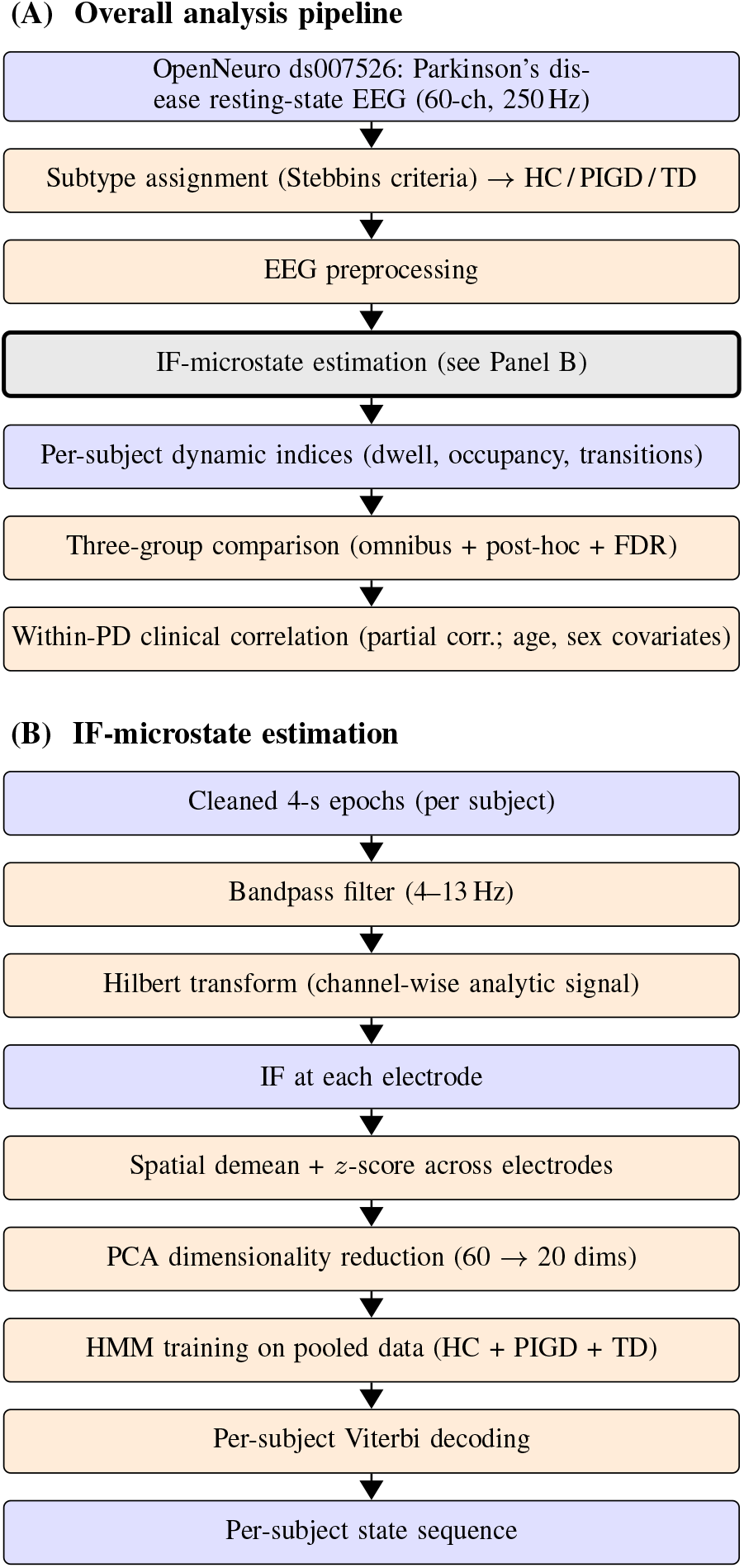
Overall analysis pipeline. (A) Macro workflow on the OpenNeuro ds007526 Parkinson’s disease resting-state EEG dataset [25]: subtype assignment by Stebbins criteria, preprocessing, IF-microstate estimation [Panel (B)], per-subject indices, three-group comparison, and within-PD clinical correlation. (B) IF-microstate estimation: Hilbert-derived IF features are dimensionality-reduced via PCA, used to train a HMM on the pooled data, and decoded per subject by Viterbi to yield state sequences. Blue boxes: data artefacts; orange boxes: processing steps; grey box in (A): the procedure detailed in (B).

### B. Participants

We performed a secondary analysis of the publicly available OpenNeuro ds007526 dataset [25], whose primary findings are reported in [52]. The data were acquired at the Laboratory of Early Markers of Neurodegeneration, Tel Aviv Sourasky Medical Center, where the study protocol was approved by the local ethics committee and conducted in accordance with the Declaration of Helsinki, and written informed consent was obtained from all participants [52]. Because the dataset is distributed in de-identified form under a CC0 licence, the present secondary analysis required no additional ethical review. As released (version 1.0.1), the dataset comprises 144 participants: 116 patients with PD and 28 HC. PD was diagnosed according to the updated Movement Disorder Society clinical diagnostic criteria; inclusion criteria were an age of 40–90 years, a Hoehn and Yahr stage of 2 or less, and a Montreal Cognitive Assessment (MoCA) score of 21 or higher, and exclusion criteria comprised prior brain surgery, stroke, significant head injury, or other major neurological disorders. Patients were assessed during the dopaminergic ON state [52]. Only the 4-min eyes-open resting-state recordings were analyzed here.

PD patients were stratified into motor subtypes from the ratio of the tremor to the PIGD subscores of the MDS-UPDRS, both of which are provided with the dataset. We applied the classification criteria of Stebbins *et al*. [3], which adapt the original UPDRS-based phenotyping of Jankovic *et al*. [2] to the MDS-UPDRS and are the same TD/PIGD phenotyping framework adopted by the dataset’s primary study [52]. Patients with a tremor/PIGD ratio of ≥ 1.15 were classified as TD and those with a ratio of ≤ 0.9 as PIGD; patients with an intermediate ratio, or with both subscores equal to zero, were labeled indeterminate, and those lacking the required subscores were left unclassified. This procedure assigned 50 patients to the PIGD group and 47 to the TD group, while 19 patients (16 indeterminate and 3 with missing subscores) were excluded from further analysis. Together with the 28 HC, this yielded an analytic sample of 125 participants (28 HC, 50 PIGD, 47 TD); no further participants were lost at the subsequent epoch-selection stage (Section III-C).

The patient-level clinical measures comprised the motor-examination (Part III) and total MDS-UPDRS scores, the MoCA, the Color Trails Test (CTT), disease duration, and the levodopa-equivalent daily dose (LEDD). Group characteristics are summarized in Table I. The three groups differed in age (Kruskal–Wallis *p* = 1.2 × 10^−3^), with HC being younger than the PD groups, and in global cognition (MoCA, *p* = 2.4 × 10^−4^); sex distribution did not differ across groups (*χ*^2^ test, *p* = 0.32). As expected from the subtyping definition, motor severity (MDS-UPDRS Part III) was higher in the PIGD than in the TD group (*p* = 1.3 × 10^−3^), whereas disease duration (*p* = 0.76) and the LEDD (*p* = 0.07) did not differ between the two PD subtypes. Age and sex were therefore included as covariates in the within-PD correlation analyses (Section III-F).

**TABLE I.** Demographic and clinical characteristics of the analytic sample (MEAN v SD). Disease duration is in months and LEDD in milligrams; both apply to PD groups only.

| | HC ($n = 28$) | PIGD ($n = 50$) | TD ($n = 47$) |
| --- | --- | --- | --- |
| Age (years) | 60.8 ± 7.5 | 67.8 ± 9.1 | 65.2 ± 9.5 |
| Sex (M/F) | 13/15 | 32/18 | 27/20 |
| MoCA | 27.4 ± 2.0 | 24.6 ± 3.3 | 26.1 ± 2.7 |
| MDS-UPDRS III | 1.4 ± 1.7 | 27.4 ± 9.7 | 21.4 ± 13.5 |
| MDS-UPDRS total | 3.3 ± 3.3 | 45.5 ± 17.3 | 33.9 ± 21.9 |
| Disease dur. (mo.) | — | 59.7 ± 46.3 | 57.2 ± 48.9 |
| LEDD (mg) | — | 479 ± 386 | 355 ± 363 |

### C. EEG Recording and Preprocessing

Resting-state EEG had been recorded from 64 scalp electrodes arranged according to the international 10–20 system while participants sat with their eyes open for approximately 4 min [25], [52]; the acquisition used a Geodesic EEG System 400 (Electrical Geodesics, Inc., Eugene, OR, USA) as documented in the dataset descriptor [25]. The recordings are distributed in the Brain Imaging Data Structure (BIDS) format at a sampling rate of 250 Hz [25], and only the resting-state runs were analyzed in the present study.

Preprocessing was performed in Python with the MNE-Python software package [53] using a fully automated, per-subject pipeline. For each recording, the four electrooculogram channels were designated as EOG, the vertex reference channel was discarded, and the electrode montage was assigned from the accompanying electrode-position file, leaving 60 scalp EEG channels. A 50-Hz notch filter and a 1–45-Hz finite-impulse-response band-pass filter were then applied. Noisy channels were identified with the PyPREP implementation of the PREP pipeline [54] and reconstructed by spherical-spline interpolation. Following the convention of the original IF-microstate framework [20], [21], no common-average re-referencing was applied, so that the recording reference was retained. Ocular and other non-neural artefacts were removed by extended-Infomax independent component analysis [55] (up to 32 components): components whose time courses correlated with the EOG channels, together with those that the ICLabel classifier [56] did not label as “brain” or “other,” were rejected before back-projection. The cleaned continuous data were segmented into non-overlapping 4-s epochs, and residual high-amplitude epochs were repaired or rejected automatically with AutoReject [57]. This procedure yielded, for each participant, a set of cleaned 4-s resting-state epochs (60 channels, 250 Hz) that served as the input to the IF-microstate analysis.

### D. IF Microstates

#### 1) Extraction of Instantaneous Frequency

In this IF-microstate approach, brain states were defined from the spatial distribution of the IF. For each participant, the longest run of consecutive clean 4-s epochs was located, and the first ten consecutive epochs were concatenated into a single 40-s segment (10,000 samples at 250 Hz); participants with fewer than ten consecutive clean epochs would have been excluded, although none met this criterion. Each channel of this segment was *z*-scored over time and then band-pass filtered between 4 and 13 Hz with a zero-phase finite-impulse-response filter, retaining both the theta (4–8 Hz) and alpha (8–13 Hz) bands [20], which are associated with large-scale neural interactions.

For each channel *i*, the analytic signal *a*_*i*_(*t*) was obtained by applying a Hilbert transform to the bandpass-filtered signal *x*_*i*_(*t*). The wrapped instantaneous phase was then defined as:

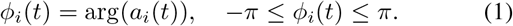

To compute IF, the phase was first unwrapped:

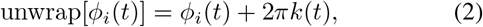

where *k*(*t*) ∈ ℤ is an integer that corrects phase discontinuities. The instantaneous frequency was calculated as the temporal derivative of the unwrapped phase.

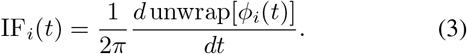

To reduce the influence of phase slips and high-frequency noise, IF_*i*_(*t*) was smoothed with a 21-sample median filter (approximately 84 ms at 250 Hz), following a previous study [17].

#### 2) Estimation of IF Microstates

To extract spatial features, at each time point the mean across electrodes was subtracted from the IF values, and the resulting deviations were *z*-score normalized across electrodes. This produced a spatially normalized 60-dimensional IF pattern at each time point [Fig. 1(B)]. To obtain a compact representation, the spatially normalized IF patterns from all participants of the three groups (HC, PIGD, and TD) were concatenated over time, and principal component analysis (PCA) was applied to reduce the 60 electrode dimensions to the leading 20 principal components.

An HMM was then fitted to the pooled, PCA-reduced IF features of all three groups to obtain a single common set of states, ensuring a consistent state identity across groups rather than fitting a separate model per group [20], [21]; each state corresponds to a recurring IF spatial pattern and is referred to as an IF microstate. We used a Gaussian HMM with full covariance matrices, whose parameters were fitted by the Baum–Welch algorithm—the expectation–maximization (EM) algorithm specialized to HMMs [58]—as implemented in the hmmlearn library [59]. The number of states *K* was treated as a model-selection problem. Candidate models with *K* ∈ {3, 4, 5, 6, 7} were compared using criteria that do not depend on the between-group contrasts of interest—the Bayesian information criterion, the balance of state occupancy, and the number of EM iterations required for convergence—with the selected value reported in Section IV (see also Supplementary Information, Fig. S1 and Table S1). For the selected *K*, individual state sequences were inferred for each participant by Viterbi decoding, from which the dynamic indices defined in Section III-E were computed.

### E. Evaluation Indices

To quantify the temporal dynamics of each IF microstate, we computed three indices: the mean dwell time, the occupancy, and the state transition probability. The mean dwell time (s) was defined as the average duration of individual microstate episodes *i*:

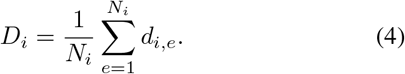

where *N*_*i*_ is the total number of times microstate *i* appears and *d*_*i,e*_ denotes the duration of the *e*-th episode of microstate *i*. Occupancy was defined as

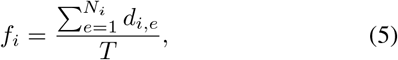

where *T* denotes the total duration of the evaluation.

Furthermore, to evaluate the transitions between the IF microstates, we computed the state transition probability for each ordered pair of microstates (*i* → *j*). The transition probability from microstate *i* to *j* was defined as follows:

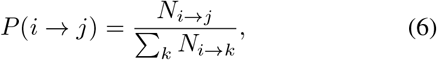

where *N*_*i*→*j*_ is the number of times a state *i* is immediately followed by a state *j*, and the denominator sums the transitions from *i* to all the possible subsequent states *k*.

### F. Statistical Analysis

For each participant and each IF microstate, the mean dwell time, the occupancy, and the between-state transition probabilities were obtained from the estimated state sequences. To reduce skewness and the influence of outliers, dwell time and occupancy were log-transformed before analysis.

The principal analysis compared the three groups (HC, PIGD, and TD) separately for each state—or each ordered state pair, in the case of transition probabilities. For each such comparison, the normality and the homogeneity of variances of the log-transformed values were assessed with the D’Agostino– Pearson and Levene tests, respectively. When both assumptions were satisfied, between-group differences were tested with a one-way Welch analysis of variance (ANOVA), which is robust to unequal variances; otherwise, the non-parametric Kruskal–Wallis test was used. Omnibus *p*-values were corrected for multiple comparisons with the Benjamini–Hochberg false-discovery-rate (FDR) procedure—across the states for dwell time and occupancy, and across all ordered state-to-state transition pairs for transition probabilities—with significance set at *q <* 0.05. Significant omnibus effects were followed by post-hoc pairwise comparisons using either the Games–Howell test (parametric), which intrinsically controls the pairwise family-wise error rate (evaluated at *α* = 0.05), or pairwise Mann–Whitney *U* tests with Benjamini–Hochberg FDR correction (non-parametric, *q <* 0.05). The Hedges’ *g* effect size is reported for each pairwise contrast, including the direct PIGD-versus-TD comparison of the two motor subtypes. Finally, to relate microstate dynamics to clinical status, partial Spearman correlations were computed within the PD group between the microstate metrics and the clinical scores (MDSUPDRS Part III and total, MoCA, CTT, disease duration, and LEDD), controlling for age and sex. The primary correlations used the two emergence metrics (dwell time and occupancy of each state); as a completeness check, the between-state transition probabilities were correlated in the same manner. These correlations were restricted to the PD group, as a correlation across the full sample would largely reflect the systematic differences between HC and PD rather than a graded relationship with disease severity. FDR correction was applied within each clinical score—across the state × metric pairs for the emergence metrics and across the state-to-state transitions for the transition probabilities. All statistical analyses were performed in Python with the Pingouin [60], SciPy, and statsmodels packages.

## IV. Results

### A. IF-Microstate Dynamics and Group Differences

We set the number of IF microstates to *K* = 3. Sweeping the candidate range *K* ∈ {3, 4, 5, 6, 7}, *K* = 3 was the smallest model that combined balanced, non-degenerate state occupancy—each state occupying between 32 % and 34 % of the time—with rapid and reliable convergence of the HMM fit; larger models produced increasingly imbalanced, low-occupancy states and, for *K* = 5 and *K* = 6, failed to converge within the iteration budget, even though the Bayesian information criterion continued to decrease with *K*. The full model-selection diagnostics are reported in the Supplementary Information (Fig. S1 and Table S1).

Fitting the three-state HMM to the pooled data yielded three IF microstates whose group-averaged topographies were posterior-weighted variants rather than qualitatively distinct maps [Fig. 2(A)]: State 1 exhibited a centro-parietal IF maximum, State 2 a bilateral posterior-temporal distribution, and State 3 an occipital-dominant maximum. Because the states were estimated from the pooled data, their topographies were, by construction, common across groups; the between-group analyses therefore concerned the temporal deployment of these shared states rather than their spatial form. Representative state sequences for one participant per group are shown in Fig. 2(B). Group differences were concentrated in State 3 [Fig. 2(C)]. The mean dwell time in State 3 increased monotonically from HC to TD to PIGD (Kruskal–Wallis *H* = 18.4, *p* = 1.0 × 10^−4^, *q* = 3.1 × 10^−4^), and all three pairwise contrasts survived FDR correction: HC versus PIGD (Hedges’ *g* = 1.06, *q* = 5.2 × 10^−5^), HC versus TD (*g* = 0.59, *q* = 0.025), and PIGD versus TD (*g* = 0.40, *q* = 0.039). This graded HC *<* TD *<* PIGD ordering indicates that the occipital-dominant State 3 was sustained for progressively longer uninterrupted intervals along the motor-subtype continuum. Dwell times in States 1 and 2 did not differ across groups (*q >* 0.36).

**Fig. 2.**
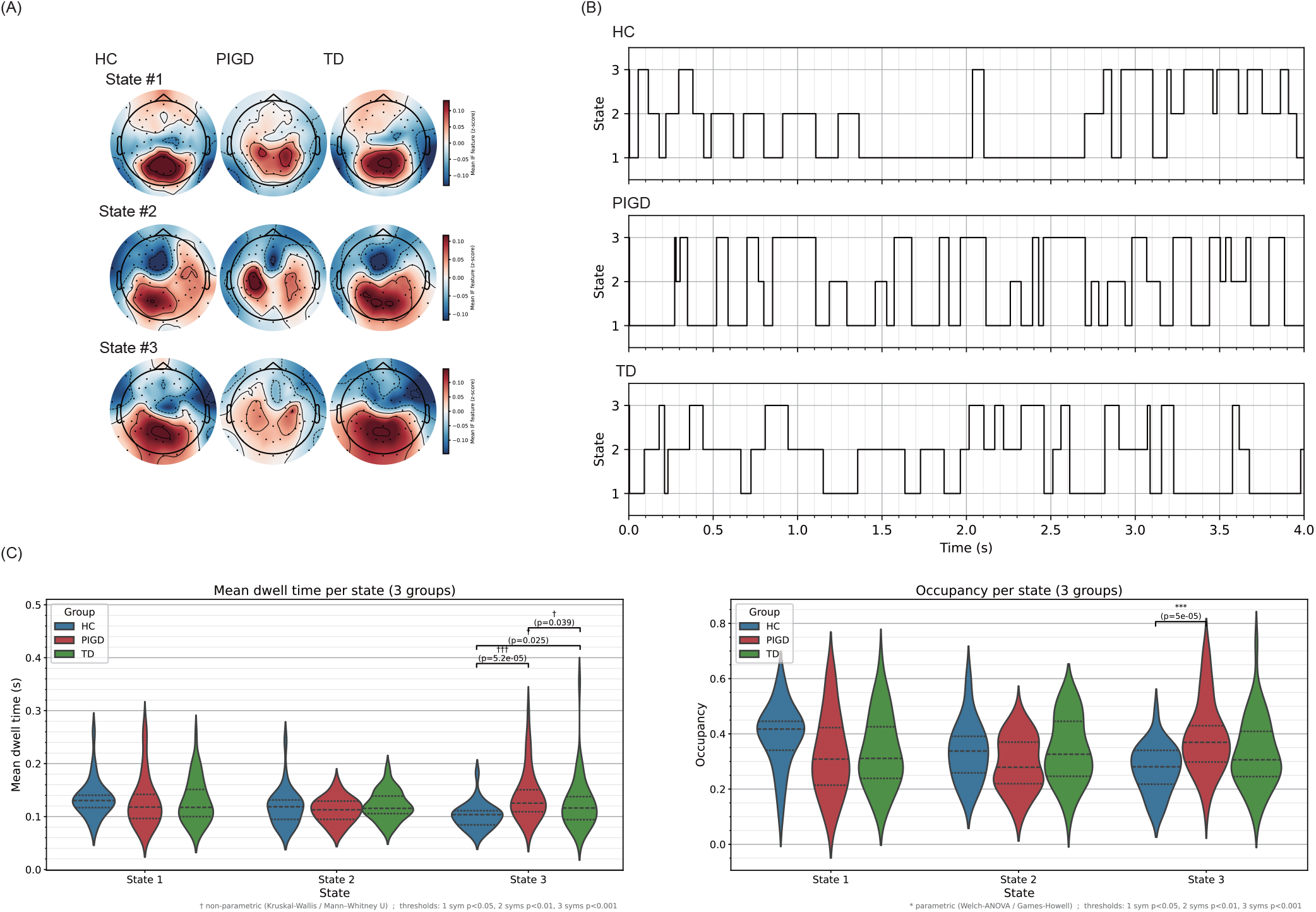
IF microstates and their group differences. (A) Group-averaged spatial topographies of the spatially normalized IF for each state (HC, PIGD, TD columns), showing posterior-weighted patterns that are common across groups. (B) Representative Viterbi state sequences for one participant per group (first 4 s). (C) Group differences in IF-microstate dynamics: (left) mean dwell time and (right) occupancy per state for HC, PIGD, and TD. Brackets denote FDR-corrected post-hoc contrasts (Games–Howell for occupancy; pairwise Mann–Whitney ***U*** for dwell time). State 3 shows a graded HC ***<*** TD ***<*** PIGD increase.

A consistent ordering was found for State 3 occupancy (one-way Welch ANOVA *F* = 10.6, *p* = 8.8 × 10^−5^, *q* = 2.7× 10^−4^; group means 0.24, 0.28, and 0.32 for HC, TD, and PIGD). Here the HC versus PIGD difference was large and significant (*g* = 0.99, *p* = 5 × 10^−5^), whereas the HC versus TD (*g* = 0.49, *p* = 0.076) and PIGD versus TD (*g* = 0.47, *p* = 0.056) contrasts were borderline. Occupancy of States 1 and 2 did not differ across groups.

The transition structure mirrored these effects (Fig. 3). Each ordered state pair was compared across the three groups with the same omnibus tests used above (Welch ANOVA or Kruskal–Wallis according to the normality check), FDR-corrected across the nine transitions, and significant omnibus effects were resolved by pairwise post-hoc tests. Every transition that differed between groups (all *q <* 0.01) involved State 3. The State 3 self-transition, which indexes state persistence, differed across the three groups (Kruskal– Wallis *H* = 18.2, *q* = 9.9 × 10^−4^) and, in the post-hoc tests, showed the same graded HC *<* TD *<* PIGD ordering as dwell time, with all three pairwise contrasts significant (HC vs PIGD *g* = 1.06; HC vs TD *g* = 0.59; PIGD vs TD *g* = 0.40; all *q <* 0.05). The remaining effects were carried chiefly by the PIGD subtype and are expressed here relative to HC: PIGD entered State 3 more frequently than HC, from both State 1 and State 2, whereas the PD groups left State 3 less frequently than HC—both PIGD and TD for the State 3 → 1 transition and PIGD for State 3 → 2. Transitions among States 1 and 2 did not differ across groups. Thus, across dwell time, occupancy, and transitions, the occipital-dominant State 3 acted as an increasingly persistent attractor— more readily entered, more strongly sustained, and less readily left—along the HC → TD → PIGD continuum, with the PIGD subtype most affected.

**Fig. 3.**
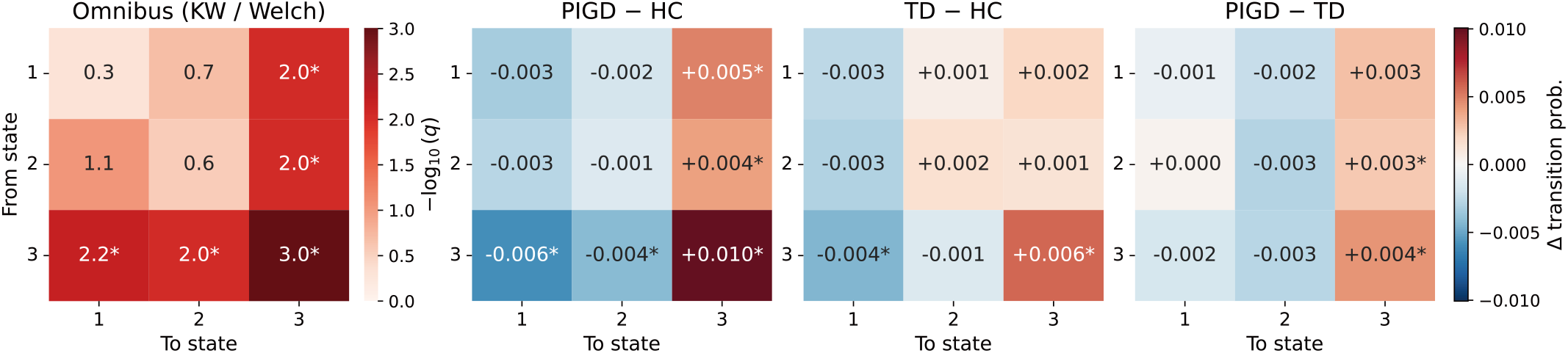
State-transition probability differences across groups. The leftmost panel shows the three-group omnibus test for each ordered state pair— Kruskal–Wallis (KW) or Welch analysis of variance (Welch), selected per cell by the normality check—as − **log**_**10**_**(*q*)**, FDR-corrected across the nine transitions; cells with ***q <* 0.05** are starred (*). The remaining three panels show the difference between two groups’ mean transition matrices (PIGD − HC, TD − HC, and PIGD − TD; rows, from-state; columns, to-state; warm colours indicate a higher probability in the first group of the pair), with a cell starred (*) when that pairwise post-hoc contrast is significant and the omnibus survived FDR (***q <* 0.05**). Differences are confined to State 3, which PIGD enters more and leaves less than HC, and whose self-transition rises along the HC ***<*** TD ***<*** PIGD continuum.

### B. Association with Clinical Measures

To test whether the IF-microstate dynamics tracked clinical status within the PD cohort, the dwell time and occupancy of each state—the two primary emergence metrics—were related to the MDS-UPDRS Part III and total scores, the MoCA, the CTT, disease duration, and the LEDD by partial Spearman correlations controlling for age and sex (Section III-F). None of the 36 resulting associations survived FDR correction within any clinical score (all *q* ≥ 0.35; *n* = 93–97), and the strongest individual trend—a negative association between State 3 dwell time and disease duration—did not reach significance even before correction (*ρ* = −0.19, *p* = 0.058). The between-state transition probabilities, examined in the same way, were likewise unrelated to any clinical score after correction (all *q* ≥ 0.26). Thus, although the State 3 dynamics separated the diagnostic groups, they did not scale with continuous measures of motor severity, cognition, disease duration, or dopaminergic medication within PD. The full correlation matrices are provided in the Supplementary Information (Tables S2 and S3).

## V. Discussion

In this study, we investigated whether the temporal dynamics of IF microstates, estimated from resting-state EEG with an HMM, can distinguish the PIGD and TD motor subtypes of PD from each other and from HC. In the ds007526 cohort, three recurrent IF microstates were identified, and the dynamics of one of them—State 3—varied systematically across groups: its mean dwell time, occupancy, and self-transition probability increased monotonically from HC through TD to PIGD, while the transitions leaving it became correspondingly less frequent, so that State 3 behaved as an increasingly persistent attractor along the HC → TD → PIGD continuum. Within the PD group, however, none of these indices was related to clinical severity, disease duration, or medication dose. The dynamics therefore stratified the motor subtypes in a graded, categorical manner without tracking continuous disease severity. In the same cohort, an unsupervised, EEG-based clustering has been used to derive data-driven neurophysiological subtypes that did not themselves differ in motor severity [52]; the present results are complementary, showing that the temporal dynamics of IF microstates vary in a graded manner across the clinically defined motor subtypes—a distinction not captured by that spectral clustering.

First, we consider why the alteration followed a graded HC *<* TD *<* PIGD ordering, with the PIGD subtype most affected. The PIGD phenotype is characterized by a heavier non-motor and cognitive burden than the TD phenotype [4]– [6]. The noradrenergic system in PD undergoes multilevel reorganization—LC cell loss, compensatory firing changes in surviving neurons, altered NA release, and downstream adrenergic receptor sensitization [23], [24]—rather than a simple monotonic depletion of cortical NA [30], [31]. Because the LC-NA system is one of the principal regulators of the balance between cortical integration and segregation [22], the observed contraction of state-switching in PIGD is plausibly a correlate of this multilevel reorganization; the TD-versus-PIGD divergence may then reflect differences in the degree or quality of neuromodulatory reorganization and its impact on large-scale network flexibility. The cross-sectional nature of the publicly available dataset used in this study [25] means this correspondence is correlational and cannot establish causality; it nonetheless aligns with the view that IF-microstate dynamics index subtype-related cortical neuromodulatory reorganization.

Second, we consider what the increased persistence of State 3 implies for cortical dynamics in PD. The combination of longer dwell times, a higher self-transition probability, and fewer outgoing transitions indicates that the cortical dynamics became progressively more likely to remain in, and less able to escape from, a single recurrent state—that is, the repertoire of state switching contracted and the dynamics grew less flexible. Flexible, wide-ranging transitions among recurring states are thought to support healthy cortical function, and a reduction of such dynamic flexibility has been reported in healthy aging and in age-related neurodegenerative disorders [20], [21]. The present results extend this picture to PD and, more specifically, indicate that the contraction of dynamic flexibility scales with motor-subtype severity, being greatest in PIGD.

Third, we consider why the microstate dynamics stratified the groups yet did not track clinical severity within PD. Two, not mutually exclusive, explanations are plausible. The dynamics may reflect a categorical, subtype-specific feature of cortical organization— plausibly the multilevel neuromodulatory reorganization in PD [23], [24]—rather than a continuous severity dimension, in which case they would separate PIGD, TD, and HC without varying monotonically with motor scores that emphasize specific motor signs. Alternatively, the within-PD analysis may have been limited in power by the restricted range of severity in this mild-to-moderate cohort (Hoehn and Yahr ≤ 2) and by the modest subgroup sizes. Either way, the dissociation indicates that the marker is not simply a proxy for overall disease severity, even if a milder severity-related modulation cannot be excluded. In relation to the biomarker roles outlined in the Introduction, the categorical separation of the groups supports a potential role in motor-subtype stratification, whereas the second anticipated role—detection of pre-motor neurodegeneration in at-risk cohorts—lies beyond the present, clinically diagnosed sample and remains to be tested.

Several limitations should be considered when interpreting these findings. First, the observed microstates were posterior-weighted variants rather than qualitatively distinct maps— a pattern also obtained with the IF-microstate framework in healthy aging [21]—so the specific topography of any individual state should not be over-interpreted. Resolving finer spatial structure would require source-level approaches, such as magnetoencephalography or EEG source localization, that identify the neural generators of the IF distribution more precisely. Second, the study was cross-sectional and based on a single cohort; longitudinal designs and replication in independent samples will be needed to establish the stability and generalizability of the subtype gradient. Third, no direct measure of any of the constituent levels of cortical noradrenergic reorganization—LC structural integrity, NA release, or adrenergic receptor density—was available in the dataset, so the proposed link to multilevel LC-NA reorganization remains hypothetical. Because LC structural integrity and cortical NA function can dissociate through compensatory firing, release, and receptor changes [23], [24], [30], no single modality could on its own adjudicate which constituent process is most closely tracked by the present EEG-derived dynamics. Multimodal studies combining IF-microstate EEG with LC neuromelanin-sensitive MRI, *α*_2_-adrenergic receptor PET, and noradrenergic biomarkers in future work would allow these relationships to be tested directly. Finally, the within-PD correlation analysis was constrained by the available sample size, limiting its power to detect associations between the microstate dynamics and clinical measures; validation in a larger cohort will be required to establish whether such associations exist.

## VI. Conclusion

Using an IF-microstate–HMM analysis of resting-state EEG, we showed that the temporal dynamics of cortical microstates differ in a graded manner across the motor subtypes of PD: an occipital-dominant state became an increasingly persistent attractor along the HC → TD → PIGD continuum, most markedly in PIGD. These dynamic differences distinguished the clinically defined subtypes even though they did not scale with continuous disease severity within PD, indicating that IF-microstate dynamics capture subtype-related differences in cortical organization and network flexibility rather than serving as a simple index of disease severity. The approach thus provides a candidate, objective EEG marker for motor-subtype stratification; realizing its translational value will require validation in larger, longitudinal cohorts.

## Supporting information

Supplemental Information: Instantaneous-Frequency EEG Microstate Dynamics Stratify Motor Subtypes in Parkinson's Disease

## Data Availability

This work is a secondary analysis of publicly available data. The resting-state EEG recordings analyzed here are openly available from the OpenNeuro repository under accession ds007526 [25]. The analysis code is available from the corresponding author upon reasonable request.

## Author Contributions

S.N. conceived and designed the study, implemented the analysis pipeline, performed the statistical analyses, and wrote the manuscript. H.W. contributed clinical and neuroscientific expertise, advised on interpretation of the findings, and critically revised the manuscript.

## Generative AI Statement

Generative AI (Claude Opus 4.7, Anthropic) was used for language refinement and to assist with development of the analysis code. The authors verified all AI-assisted output and take full responsibility for the content of this article.

