## Supplemental Information: Instantaneous-Frequency EEG Microstate Dynamics Stratify Motor Subtypes in Parkinson's Disease for "Instantaneous-Frequency EEG Microstate Dynamics Stratify Motor Subtypes in Parkinson’s Disease"

Sou Nobukawa, *Senior Member, IEEE*, and Hirohisa Watanabe

### I. Selection of the Number of HMM States

The number of states $K$ of the hidden Markov model (HMM) was selected by sweeping $K$ over the range $\{3, 4, 5, 6, 7\}$ and evaluating each candidate model with criteria that do not depend on the between-group contrasts of interest, so as to avoid biasing the model order towards any particular group difference: (i) the Bayesian Information Criterion (BIC), (ii) the balance of state occupancy (the minimum and maximum occupancy across states), and (iii) the convergence behavior of the expectation–maximization (EM) algorithm (the number of iterations required). Figure S1 summarizes these diagnostics across the swept values of $K$, and Table S1 reports the corresponding numerical values.

BIC decreased monotonically with $K$ and was therefore minimized by the largest model ($K = 7$); the $K = 7$ solution, however, was dominated by a marginally occupied state (minimum occupancy $\approx 8\,\%$), and models with $K \geq 5$ exhibited substantially imbalanced occupancy, with the minimum state occupancy approaching $5\,\%$—the level below which a state is occupied too rarely to be reliably estimated. The $K = 5$ and $K = 6$ fits also required the full iteration budget (500 EM iterations), whereas $K = 3$, $K = 4$, and $K = 7$ converged in fewer iterations. $K = 3$ yielded the most balanced occupancy profile (all three states between 0.318 and 0.343) and converged rapidly (114 iterations). Accordingly, $K = 3$ was selected for the main analysis as the smallest model producing balanced, non-degenerate state occupancy and a stable fit, rather than on the basis of any group-difference statistic.

### II. Clinical Correlations within the PD Group

Table S2 lists the partial Spearman correlation coefficients (controlling for age and sex) between each instantaneous-frequency (IF) microstate metric—the mean dwell time and the occupancy of States 1–3—and the clinical scores, computed within the Parkinson's-disease (PD) group. No coefficient was significant after the Benjamini–Hochberg false-discovery-rate (FDR) correction within any score (all $q \geq 0.35$; $n = 93$–$97$ PD participants, depending on score availability).

The same analysis applied to the between-state transition probabilities is reported in Table S3; none of these coefficients was significant after FDR correction within any score either (all $q \geq 0.26$).

See the main manuscript for affiliations and funding acknowledgements.

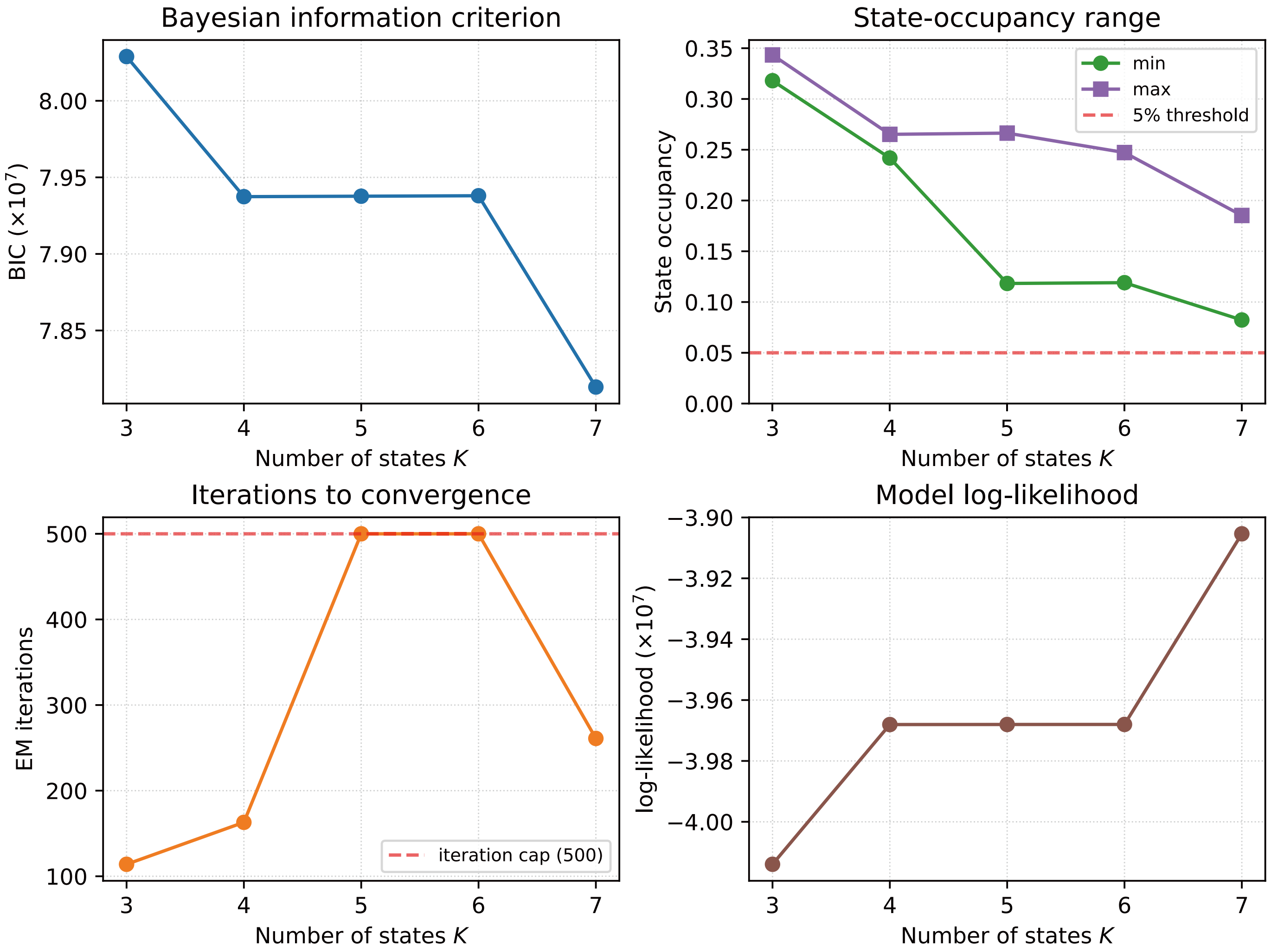


Fig. S1: HMM $K$-sweep diagnostics. Each panel summarizes one outcome-independent diagnostic across candidate state numbers $K \in \{3, 4, 5, 6, 7\}$. Top-left: Bayesian Information Criterion (BIC; lower indicates a better fit–complexity trade-off). Top-right: minimum and maximum state occupancy (5 % threshold shown as a dashed line). Bottom-left: number of EM iterations to convergence (dashed line: the 500-iteration cap). Bottom-right: model log-likelihood. $K = 3$ was selected as the smallest model with balanced, non-degenerate state occupancy and a rapidly converging fit.

TABLE S1: HMM $K$-sweep summary statistics. BIC, AIC, log-likelihood, model size, minimum/maximum occupancy, and the full per-state occupancy vector are reported for each candidate number of states $K$, together with the number of EM iterations. The EM algorithm was capped at 500 iterations; the models that reached this cap ($K = 5$ and $K = 6$) did not converge within the budget.

| $K$ | log-lik | BIC | AIC | $n_{\text{params}}$ | min occ. | max occ. | Per-state occupancy | EM iter. |
|---|---|---|---|---|---|---|---|---|
| 3 | $-4.014 \times 10^7$ | $8.029 \times 10^7$ | $8.028 \times 10^7$ | 698 | 0.318 | 0.343 | $0.343, 0.318, 0.339$ | 114 |
| 4 | $-3.968 \times 10^7$ | $7.937 \times 10^7$ | $7.936 \times 10^7$ | 935 | 0.242 | 0.265 | $0.265, 0.242, 0.247, 0.246$ | 163 |
| 5 | $-3.968 \times 10^7$ | $7.938 \times 10^7$ | $7.936 \times 10^7$ | 1174 | 0.118 | 0.266 | $0.248, 0.118, 0.266, 0.243, 0.125$ | 500 |
| 6 | $-3.968 \times 10^7$ | $7.938 \times 10^7$ | $7.936 \times 10^7$ | 1415 | 0.119 | 0.247 | $0.247, 0.133, 0.247, 0.122, 0.119, 0.133$ | 500 |
| 7 | $-3.905 \times 10^7$ | $7.813 \times 10^7$ | $7.811 \times 10^7$ | 1658 | 0.082 | 0.185 | $0.082, 0.142, 0.183, 0.084, 0.160, 0.164, 0.185$ | 261 |

*Abbreviation:* AIC, Akaike Information Criterion.

TABLE S2: Partial Spearman correlations ($\rho$, controlling for age and sex) between IF-microstate metrics and clinical scores, computed within the PD group. None survived FDR correction within any score (all $q \geq 0.35$; $n = 93$–$97$ participants).

| | State 1 | | State 2 | | State 3 | |
|---|---|---|---|---|---|---|
| Clinical score | Dwell | Occ. | Dwell | Occ. | Dwell | Occ. |
| MDS-UPDRS Part III | −0.03 | −0.08 | −0.14 | −0.13 | 0.09 | 0.13 |
| MDS-UPDRS total | −0.09 | −0.11 | −0.13 | −0.10 | 0.11 | 0.16 |
| MoCA | 0.00 | 0.00 | −0.03 | −0.08 | 0.08 | 0.07 |
| CTT | 0.04 | 0.06 | 0.09 | 0.09 | −0.10 | −0.15 |
| Disease duration | 0.02 | 0.09 | −0.16 | −0.07 | −0.19 | −0.11 |
| LEDD | 0.04 | 0.07 | −0.10 | −0.05 | −0.11 | −0.10 |

*Abbreviations:* FDR, false discovery rate; MDS-UPDRS, Movement Disorder Society Unified Parkinson's Disease Rating Scale (Part III and total); MoCA, Montreal Cognitive Assessment; CTT, Color Trails Test; LEDD, levodopa equivalent daily dose.

TABLE S3: Partial Spearman correlations ($\rho$, controlling for age and sex) between IF-microstate transition probabilities (State $i \rightarrow$ State $j$) and clinical scores, computed within the PD group. None survived FDR correction within any score (all $q \geq 0.26$; $n = 93$–$97$ participants). Clinical-score abbreviations as in Table S2.

| Transition | UPDRS-III | UPDRS-tot. | MoCA | CTT | Dis. dur. | LEDD |
|---|---|---|---|---|---|---|
| State 1 → State 1 | −0.02 | −0.09 | 0.00 | 0.04 | 0.02 | 0.04 |
| State 1 → State 2 | −0.12 | −0.05 | −0.02 | 0.01 | −0.02 | −0.03 |
| State 1 → State 3 | 0.18 | 0.20 | 0.00 | −0.06 | −0.05 | −0.05 |
| State 2 → State 1 | −0.02 | −0.02 | −0.01 | 0.08 | 0.19 | 0.12 |
| State 2 → State 2 | −0.14 | −0.13 | −0.04 | 0.10 | −0.16 | −0.10 |
| State 2 → State 3 | 0.14 | 0.12 | 0.05 | −0.21 | 0.02 | −0.05 |
| State 3 → State 1 | −0.05 | −0.07 | −0.02 | 0.10 | 0.17 | 0.09 |
| State 3 → State 2 | −0.13 | −0.10 | −0.10 | 0.08 | 0.09 | 0.05 |
| State 3 → State 3 | 0.10 | 0.11 | 0.08 | −0.10 | −0.19 | −0.11 |
